# Daily exposure to PM_2.5_ and 1.5 million deaths: A time-stratified case-crossover analysis in the Mexico City Metropolitan Area

**DOI:** 10.1101/2023.01.15.23284576

**Authors:** Iván Gutiérrez-Avila, Horacio Riojas-Rodríguez, Elena Colicino, Johnathan Rush, Marcela Tamayo-Ortiz, Víctor Hugo Borja-Aburto, Allan C. Just

**Affiliations:** Department of Environmental Medicine and Public Health, Icahn School of Medicine at Mount Sinai, New York, NY, USA; Dirección de Salud Ambiental, Instituto Nacional de Salud Pública, Cuernavaca Morelos, México; Instituto Mexicano del Seguro Social. Unidad de Investigación en Salud Ocupacional, México City, México; Instituto Mexicano del Seguro Social, México City, México; Institute for Exposomic Research, Icahn School of Medicine at Mount Sinai, New York, NY, USA

**Keywords:** PM_2.5_, cause-specific mortality, short-term exposure, case-crossover study

## Abstract

**Background:** Satellite-based PM_2.5_ predictions are being used to advance exposure science and air-pollution epidemiology in developed countries; including emerging evidence about the impacts of PM_2.5_ on acute health outcomes beyond the cardiovascular and respiratory systems, and the potential modifying effects from individual-level factors in these associations. Research on these topics is lacking in Latin America.

**Methods:** We used a time-stratified case-crossover study design with 1,479,950 non-accidental deaths from Mexico City Metropolitan Area for the period of 2004-2019. Daily 1x1 km PM_2.5_ (median=23.4 μg/m^3^; IQR=13.6 μg/m^3^) estimates from our satellite-based regional model were employed for exposure assessment at the sub-municipality level. Associations between PM_2.5_ with broad-category (organ-system) and cause-specific mortality outcomes were estimated with distributed lag conditional logistic models. We also fit models stratifying by potential individual-level effect modifiers including; age, sex, and individual SES-related characteristics namely: education, health insurance coverage, and job categories.

**Results:** PM_2.5_ exposure was associated with higher total non-accidental, cardiovascular, cerebrovascular, respiratory, and digestive mortality. A 10-μg/m^3^ PM_2.5_ higher cumulative exposure over one week (lag_06_) was associated with higher cause-specific mortality outcomes including hypertensive disease [2.28% (95%CI: 0.26%–4.33%)], acute ischemic heart disease [1.61% (95%CI: 0.59%–2.64%)], other forms of heart disease [2.39% (95%CI: -0.35%–5.20%)], hemorrhagic stroke [3.63% (95%CI: 0.79%–6.55%)], influenza and pneumonia [4.91% (95%CI: 2.84%–7.02%)], chronic respiratory disease [2.49% (95%CI: 0.71%–4.31%)], diseases of the liver [1.85% (95%CI: 0.31%–3.41%)], and renal failure [3.48% (95%CI: 0.79%–6.24%)]. No differences in effect size of associations were observed between SES strata.

**Conclusions:** Exposure to PM_2.5_ was associated with mortality outcomes beyond the cardiovascular and respiratory systems, including specific death-causes from the digestive and genitourinary systems, with no indications of effect modification by individual SES-related characteristics.

## Background

Human exposure to fine particulate matter (PM_2.5_) is a well-documented risk factor for cardiovascular and respiratory mortality and morbidity. Nonetheless, exposure to PM_2.5_ can affect nearly every organ system through different mechanisms of damage, including oxidative stress, systemic inflammation, and immune dysregulation (Keswani Anjeni et al., 2022). Recent epidemiologic studies have reported associations between short- and long-term exposure to PM_2.5_ with mental, behavioral, nervous, cerebrovascular, metabolic, digestive, and genitourinary mortality and morbidity, mostly in the US and Asia (Bowe et al., 2019; Li et al., 2018; Wang et al., 2020; Yu et al., 2019). Little evidence on acute mortality outcomes beyond the cardiovascular and respiratory spectrum have been reported in countries from low- and middle-income regions, including Latin America. Research focused on associations from under-studied causes of death is important to inform concentration-response functions that may be used in health impact assessments, including those quantifying the benefits of improving air quality. In the public-policy framework, omission of some health impacts may lead to underestimation of the benefits from interventions aimed to reduce the burden of disease from PM_2.5_ relative to their costs (World Health Organization, 2013).

The development of highly spatially-resolved PM_2.5_ models (1x1-km PM_2.5_ predictions) based on satellite data, has facilitated the assessment of human exposure at fine spatial scales, even in regions with low to moderate PM_2.5_ monitoring, such as Latin America (Gouveia et al., 2021). Also, the use of satellite-based PM_2.5_ models’ predictions, as well as the combinations of different methods to predict PM_2.5_ for epidemiologic research, may reduce exposure measurement error, compared to the use of city-wide averages from small ground monitoring networks (Alexeeff et al., 2015; Samoli et al., 2020; Wei et al., 2022). Highly spatially-resolved PM_2.5_ predictions for exposure assessment along with individual-level health records, allows exploration of effect modification by the inclusion of individual (or area-level shared) characteristics which are difficult to consider when using aggregated data, like in traditional city-wide time-series studies (Wei et al., 2019; Yitshak-Sade et al., 2019). Age, sex, socio-economic status (SES), and race, are among the factors that seem to modify vulnerability to PM_2.5_ exposure in high-income countries (Franklin et al., 2006; Heo et al., 2022; Renzi et al., 2021; Wang et al., 2020). Such characteristics are of interest for identification of vulnerable populations, science-based policy and targeted public-health interventions (Xu et al., 2020). However, little has been reported about the potential modifying effects of these factors in middle-income countries like Mexico, where their distribution may differ compared to other regions.

In this study, we analyzed all ∼1.5 million adult mortality records from the Mexico City Metropolitan Area for the period from 2004 to 2019, and estimated the daily percent increase in non-accidental mortality for broad-category and cause-specific mortality outcomes; as well as sex and age-group (adults and the elderly) categories, and SES-related indicators, associated with a 10-μg/m^3^ higher PM_2.5_ concentration.

## Methods

### Mortality data

We obtained mortality records from the National Institute of Statistics and Geography of Mexico (INEGI) for the period spanning February 1^st^ 2004 to December 31^st^, 2019. We excluded the month of January 2004 in order to keep only case-days and control-days with complete exposure histories of PM_2.5_ and temperature, i.e. up to one week (lag 6) before the date of event. All mortality records included information on date of death, geographic identifiers for place of residence, underlying cause of death classified according to the International Classification of Diseases Tenth Revision (ICD-10), sex, age, education, healthcare affiliation, and job category.

We restricted our analysis to non-accidental causes of death excluding most ICD-10 codes from the blocks V–Y (i.e. various accidents and side effects of treatments), and deaths in people ≥ 18 years-old. ICD-10 codes X6-Y0 were retained based on the evidence that PM_2.5_ can trigger intentional self-harm and aggressive behavior (Berman et al., 2019; Davoudi et al., 2021). We organized individual ICD-10 coded death records into mutually exclusive broad-category (organ-system) and cause-specific mortality outcomes. The selected ICD-10 codes included in our research were chosen to facilitate comparison with recently-published studies focused on under-studied associations between short-term exposure to PM_2.5_ with mortality and morbidity outcomes (Li et al., 2018; Yu et al., 2019).

To address the potential modifying effects of age, sex and SES on the PM_2.5_-mortality relationship, we performed stratified analyses by age-group (adults: 18-64 years-old, and elderly: +65 years-old) and sex (males and females) for all broad-category mortality outcomes. For all non-accidental mortality, we also performed stratified analyses by age-group (adults and the elderly), degree of education (basic and more than basic education level), type of healthcare affiliation (having health social security affiliation or not), and employment status based on job categories. We used health social security affiliation rather than health insurance alone, since it is related to formal employment and other individual social benefits that differ from specific healthcare programs aimed to provide health services to the most vulnerable populations in Mexico. Employment status was defined from the original job category variable in the mortality records that included the categories “Does not work” and “Looking for a job”, which were combined into one single category “Not employed”. However, this categorization does not rule out the possibility that part of the category “Does not work” can also include retired people, with a higher SES compared with those who are actually unemployed. The rest of the job categories were combined into the category “Employed”. We also explored education-specific and job-specific associations. For the latter, given that the Mexican classification of occupations was updated in 2012, and specific occupations coded before 2012 were redistributed into new job categories in the most recent version, it was not possible to homogenize all job categories for the whole period of study. Thus, analyses of specific job categories were restricted to the period from 2013 to 2019. We compared if the effect size of PM_2.5_ exposure differed by age group, and also by education, healthcare affiliation, and employment status, in the broad-category mortality outcomes, and total non-external mortality by following the methods described by Altman and Bland, 2003.

### Environmental data

We utilized daily PM_2.5_ predictions with spatial resolution of 1×1 km from our recently developed model based on extreme gradient boosting (XGBoost), and inverse-distance weighting (IDW) that uses aerosol optical depth data, meteorology, and land-use variables to predict daily mean PM_2.5_ for the Mexico City Metropolitan Area (Gutiérrez-Avila et al., 2022). Daily mean air temperature with the same 1x1 km resolution was obtained from our satellite-based land surface temperature model for Central Mexico (Gutiérrez-Avila et al., 2021). We restricted our analyses to the spatial domain with available predictions for both PM_2.5_ and temperature exposures over the Mexico City Metropolitan Area. The study area was composed of 667 sub-municipal geographic areas defined as “localities” by the Instituto Nacional de Estadistica y Geografia. Localities correspond to the third level of subnational division in Mexico, after states and municipalities (Instituto Nacional de Geografía y Estadística, 2010). The total population in the Mexico City Metropolitan Area in 2010 was 20,116,842 (Secretaría de Desarrollo Agrario, Territorial y Urbano, 2018). For the same year, the population living in the localities included in our study region was 19,711,516 inhabitants, ∼96% of the total Mexico City Metropolitan Area. Urban localities had a population of 19,346,527 (median of 8,523 inhabitants; range of 910 to 1,815,786) inhabitants, and rural localities had a total population of 364,989 (median of 582 inhabitants; range 1 to 5,135) inhabitants. We assigned exposures to PM_2.5_ and temperature for all mortality records at the locality level. For 255 urban localities with census-provided polygons (median land area of 2.9 km^2^ ; range 0.2 to 129.4 km^2^), we estimated daily exposures using population-weighted aggregation with population density from the Gridded Population of the World (GPWv4) ∼1-km raster cells (Center for International Earth Science Information Network-CIESIN-Columbia University, 2016), which utilizes data from the 2010 Mexican Census, and the R package *exactextractr* (Baston, 2020). For 412 rural localities, the census only assigned points (rather than enclosing polygons), and for this subset of rural localities we assigned exposure to PM_2.5_ and temperature using the 1x1 km grid cell containing the corresponding census-assigned points.

### Statistical Analysis

We estimated the association between short-term exposure to PM_2.5_ with mortality using a time-stratified case-crossover design (Levy et al., 2001). Case days were defined as the date of death, and control days (3 to 4 days per month) were chosen on the same day of week as the case day within the same month and year. This approach controls for potential confounding effects by day of week, seasonal patterns and long-term time trends (Janes et al., 2005). Time-invariant covariates are controlled by design (within-person matching), and are not considered to be confounders; thus, daily variations in exposure to PM_2.5_ and temperature on the case day are compared to exposures on control days. All models were adjusted for non-linear effects of temperature with quadratic b-splines (4 degrees of freedom), and equally-spaced knots in the space of this predictor (Gasparrini, 2021). The odds ratios for all dependent variables associated with short-term exposure to PM_2.5_ were estimated with linear terms in stratified Cox proportional hazards models, equivalent to conditional logistic regression. We explored non-linearities in the concentration-response relationships between PM_2.5_ with all broad-category mortality outcomes by comparing the fit of generalized additive models (GAMs) including parsimonious non-linear (penalized thin-plate splines) PM_2.5_ terms (lag_0_ to lag_6_) with the fit of linear PM_2.5_ terms using likelihood ratio tests. To address the delayed effects of exposure to PM_2.5_ on all mortality outcomes we included distributed lag terms up to 6 days before the case day (seven terms for lags 0 to 6) to estimate mortality risks (Gasparrini et al., 2010). For ease of interpretation, all odds ratios were converted to percentage increase per 10 μg/m^3^ higher PM_2.5_.

All analyses were performed in R version 4.2.1 (R Core Team, 2020) with packages: *data*.*table* (Dowle et al., 2019), *survival (Therneau and Lumley, 2015), dlnm (Gasparrini, 2011), and mgcv (Wood SN, 2015)*. Because the records used in all analyses were publicly available, the PI previously received a determination of exempt human research: 45 CFR 46. 101(b) (Category 4) from the Institutional Review Board of the Icahn School of Medicine at Mount Sinai.

## Results

The total number of non-accidental deaths (≥18 years-old) analyzed from 2004 to 2019 in the Mexico City Metropolitan Area was 1,479,950. Table 1 describes the demographic characteristics of the study population.

**Table 1.** Demographic characteristics of the analyzed deaths in people >18 years-old in the Mexico City Metropolitan Area from 2004-2019.

| **Characteristic** | **Frequency** |
| --- | --- |
| All non-accidental deaths | 1,479,950 (100%) |
| **Age group** |  |
| Elderly 65+ | 927,795 (62.7%) |
| Adults 18-64 | 552,155 (37.3%) |
| **Sex** |  |
| Men | 751,804 (50.8%) |
| Women | 728,146 (49.2%) |
| **Education** |  |
| Primary School | 731,468 (49.4%) |
| Mid-High School | 340,172 (23%) |
| No education | 191,426 (12.9%) |
| College | 153,100 (10.3%) |
| Unspecified | 63,784 (4.3%) |
| **Insurance** |  |
| Health social security affiliation | 928,678 (62.8%) |
| No health social security affiliation | 496,491 (33.5%) |
| Unspecified | 54,781 (3.7%) |
| **Job type** |  |
| Does not apply before 2013 | 738,294 (49.9%) |
| Not employed | 477,573 (32.3%) |
| Sales workers | 53,918 (3.6%) |
| Unspecified | 48,332 (3.3%) |
| Craft and related trades workers | 47,834 (3.2%) |
| Professionals and technicians | 44,423 (3%) |
| Plant and machine operators | 21,727 (1.5%) |
| Agricultural, forestry and fishery | 12,926 (0.9%) |
| Personal services | 11,337 (0.8%) |
| Clerical support | 9,581 (0.6%) |
| Elementary occupations | 9,475 (0.6%) |
| Managers | 4,530 (0.3%) |
| **PM<sub>2.5</sub>** |  |
| Mean (SD) | 24.6 (10.8) |
| Median [Min, Max] | 23.4 [0.200, 202] |
| **Tmean** |  |
| Mean (SD) | 16.6 (2.58) |
| Median [Min, Max] | 16.8 [3.30, 24.1] |

There were slightly more deaths in men than women, and deaths in the elderly (≥65 years-old) accounted for ∼63% of all deaths. Around 50% of the decedents attended only elementary school, 63% had health social security affiliation, and 32% were not working at the time of death. The five most common job categories in the mortality records for the period 2013-2019 were: not-employed or not working at the time of death, sales, unspecified occupations, craft and related trades, and professionals and technicians. Over the study period, the mean population-weighted exposure to PM_2.5_ and temperature in the study region were 24.6μg/m^3^, and 16.6º C, respectively.

Table 2 shows the total number of organ-system and cause-specific deaths (≥18 years-old) analyzed over the study period. Circulatory system diseases were the most prevalent death causes, accounting for almost 30% of the total, followed by digestive (12%) and respiratory (10%) diseases.

**Table 2.** Underlying cause-specific mortality (>18 years-old) in the Mexico City Metropolitan Area from 2004-2019.

| **Disease** | **ICD code** | **Frequency** |
| --- | --- | --- |
| All non-accidental deaths |  | 1,479,950 |
| **Disease group** |  |  |
| Circulatory System | I00-I99 | 427,420 |
| Digestive system | K00-K93 | 172,906 |
| Respiratory system | J00-J99 | 142,866 |
| Genitourinary | N00-N99 | 57,372 |
| Nervous system | G00-G99 | 22,735 |
| Intentional self-harm | X60-X84 | 10,021 |
| Mental and behavioral | F00-F99 | 9,577 |
| **Specific death cause** |  |  |
| Acute ischemic heart disease | I20-I22, I24 | 212,201 |
| Diseases of the liver | K70-K77 | 92,562 |
| Chronic respiratory disease | J40-J47 | 67,062 |
| Hypertensive diseases | I10-I15 | 53,675 |
| Influenza and Pneumonia | J09-J18 | 52,972 |
| Renal failure | N17-N19 | 30,787 |
| Other forms of heart disease | I30-I52 | 29,516 |
| Stroke hemorrhagic | I60-I62 | 28,412 |
| Chronic ischemic heart disease | I25 | 21,313 |
| Disorders of gallbladder, biliary tract, and pancreas | K80-K87 | 16,266 |
| Diseases of esophagus, stomach and duodenum | K20-K31 | 12,322 |
| Stroke ischemic | I63 | 10,684 |
| Suicides | X6-X84 | 9,954 |
| Pulmonary heart disease | I26-I28 | 6,792 |
| Diseases of the arteries | I70-I79 | 6,299 |
| Chronic rheumatic heart diseases | I05-I09 | 4,314 |
| Extrapyramidal and movement disorders | G20-G26 | 3,531 |

Among the specific death causes, acute ischemic heart disease and hypertensive disease were the leading causes from the circulatory system. Diseases of the liver were the leading death causes from the digestive system, and chronic respiratory disease, influenza and pneumonia were the most common causes of death from the respiratory system (Table 2).

Results from our assessment of linearity in the concentration-response functions between short-term exposure to PM_2.5_ with all broad-category mortality outcomes showed no significant differences in model’s fit when including non-linear PM_2.5_ exposures; therefore all our further results are based on linear concentration-response functions. Figure 1 shows associations between PM_2.5_ exposure with the broad-category death causes analyzed over the study period. For consistency with previous studies, two-days (lag_0_ + lag1) and one-week, (sum from lag_0_ to lag_6_) cumulative associations with PM_2.5_ are shown. Among the broad-category death causes, two-days exposure to PM_2.5_ (lag_01_) was associated with total higher non-accidental mortality [adults = 0.72% (95%CI: 0.26%–1.19%), elderly = 1.04% (95%CI: 0.68%–1.40%)], with sex-specific associations for men of 0.84% (95%CI: 0.44%–1.24%) and women of 1.00% (95%CI: 0.60%–1.41%), cardiovascular mortality [adults = 1.03% (95%CI: -0.3%–2.10%), elderly = 1.01% (95%CI: 0.41%–1.62%)], cerebrovascular mortality [adults = 2.84% (95%CI: 0.52%– 5.21%)], respiratory mortality [adults = 2.51% (95%CI: 0.63%–4.44%), elderly = 2.02% (95%CI: 1.00%–3.04%)] and digestive mortality [elderly = 1.16% (95%CI: -0.01%–2.35%)]. Cumulative associations over one week (lag_06_) were in general larger compared with associations for lag_01_ for most mortality outcomes; with the largest associations observed for cerebrovascular [adults = 3.65% (95%CI: 0.49%–6.91%)], respiratory [adults = 2.61% (95%CI: 0.04%–5.25%), elderly = 3.46% (95%CI: 2.05%– 4.90%)], and genitourinary [adults = 3.35% (95%CI: 0.11%–6.69%), elderly = 2.30% (95%CI: -0.17%– 4.83%)] mortality. Our test to identify differences in effect sizes by age and sex in all broad-categories of death did not suggest effect modification from these variables in the associations between PM_2.5_ and mortality. Results for specific single lag (lag_0_ … lag_6_) and cumulative associations (lag_01_ … lag_06_) for all broad-categories of mortality and PM_2.5_ are reported in Table S1.

**Figure 1.**
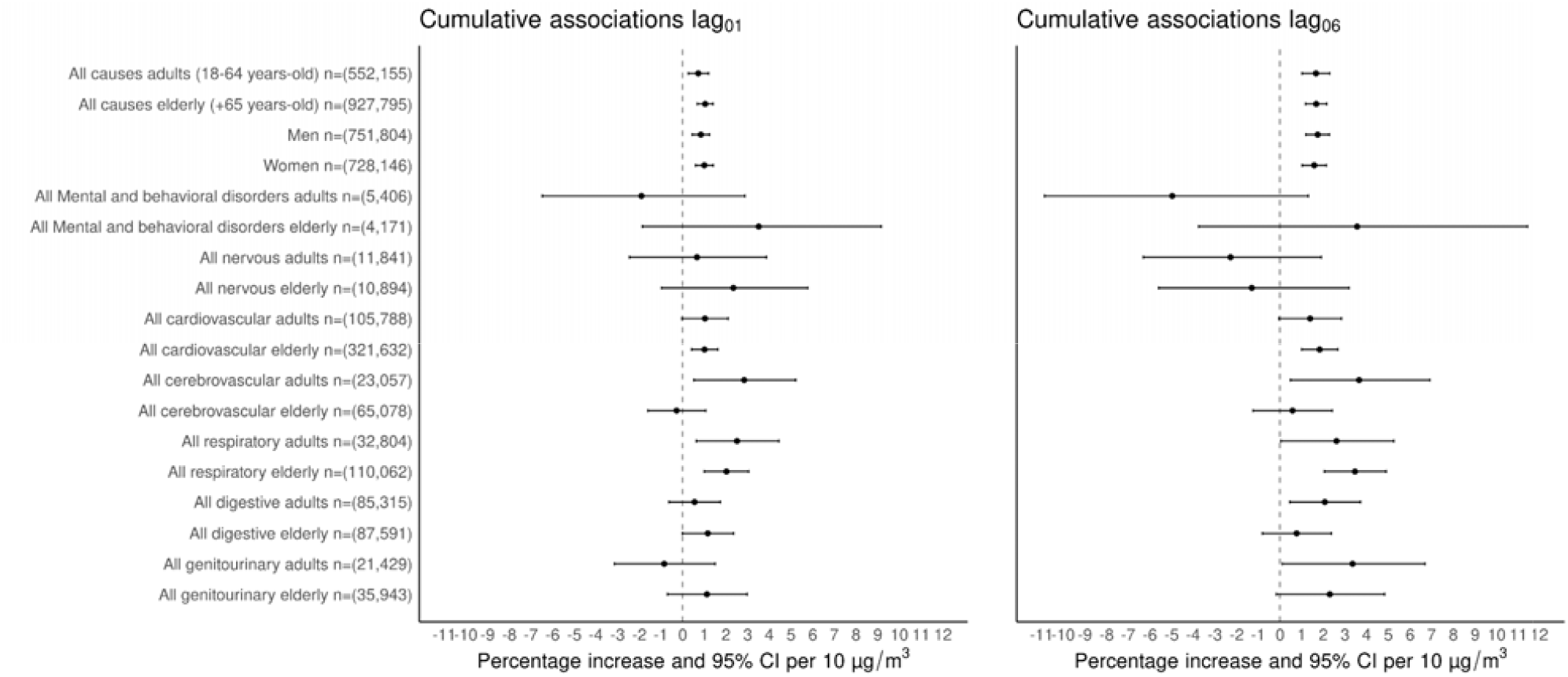
Cumulative percentage increase (%) and 95% CI for broad-category death causes by age group (adults ≥18-64 years-old and elderly ≥65 years-old) per 10 μg/m^3^ increase of PM_2.5_ over two days (lag_01_) and one week (lag_06_) in the Mexico City Metropolitan Area for 2004-2019.

Associations between PM_2.5_ exposure and cause-specific mortality for lag_01_ and lag_06_ are shown in Figure 2. Cumulative exposure to PM_2.5_ over two days (lag_01_) was associated with mortality from hypertensive disease [1.14% (95%CI: -0.33%–2.62%)], acute ischemic heart disease [1.21% (95%CI: 0.47%–1.96%)], influenza and pneumonia [2.97% (95%CI: 1.48%–4.48%)], and chronic respiratory disease [1.51% (95%CI: 0.23%–2.81%)]. Associations for cumulative exposure to PM_2.5_ over one week (lag_06_) were observed for hypertensive disease [2.28% (95%CI: 0.26%–4.33%)], acute ischemic heart disease [1.61% (95%CI: 0.59%–2.64%)], other forms of heart disease [2.39% (95%CI: -0.35%–5.20%)], hemorrhagic stroke [3.63% (95%CI: 0.79%–6.55%)], influenza and pneumonia [4.91% (95%CI: 2.84%–7.02%)], chronic respiratory disease [2.49% (95%CI: 0.71%–4.31%)], diseases of the liver [1.85% (95%CI: 0.31%–3.41%)], and renal failure [3.48% (95%CI: 0.79%–6.24%)].

**Figure 2.**
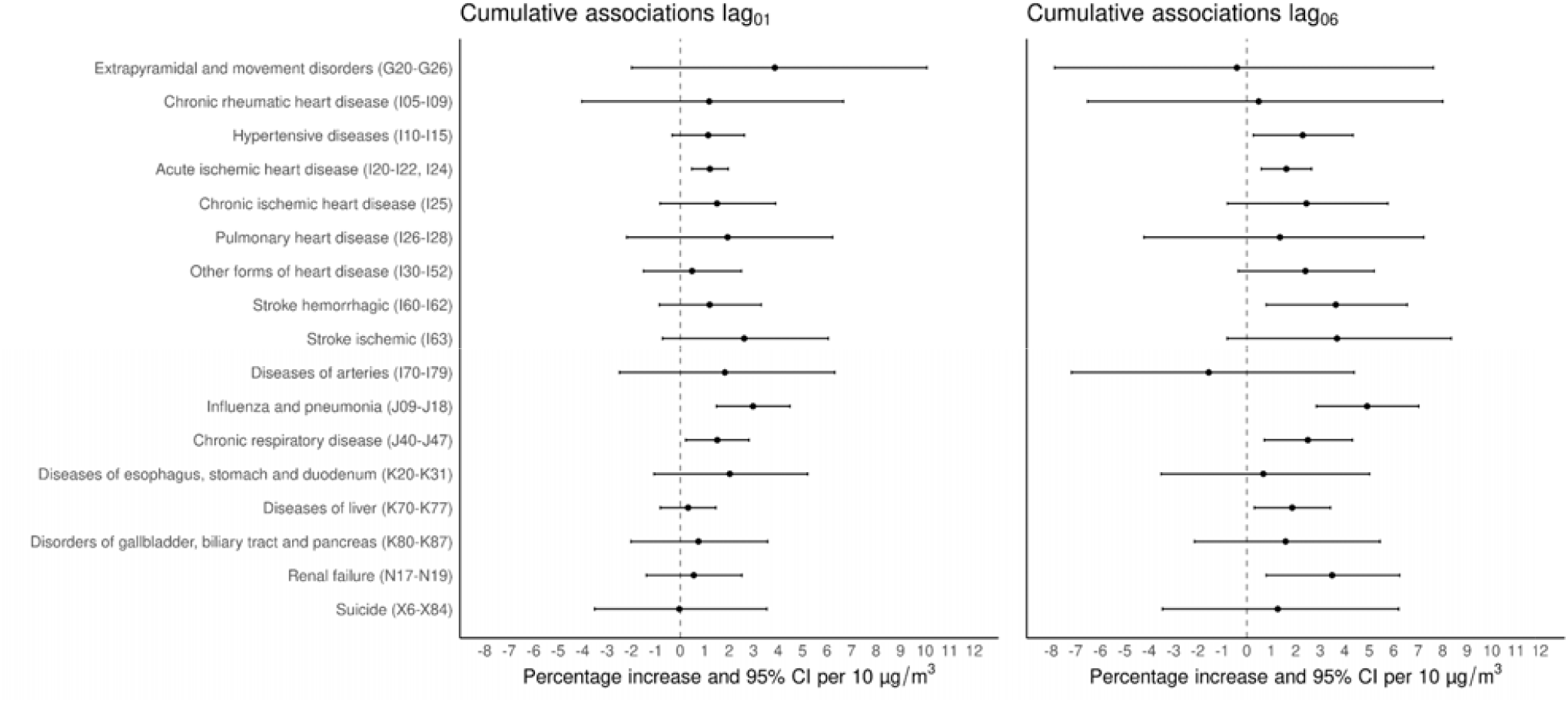
Cumulative percentage increase (%) and 95% CI for cause-specific mortality by age group (adults ≥18 years-old and elderly ≥65 years-old) per 10 μg/m^3^ increase of PM_2.5_ over two (lag_01_) days and one week (lag_06_) in the Mexico City Metropolitan Area for 2004-2019.

Cumulative associations from distributed lags can capture the impact of multiple days after initial exposure to PM_2.5_ on mortality. In general, consistent associations between short-term exposure to PM_2.5_ with all-cause (i.e. total non-accidental) or broad-category death causes (mostly from cardiovascular and respiratory diseases) have been observed during the first week of exposure across study sites. Therefore, communicating results for lag_01_ and lag_06_, has become the standard, as the evidence shows immediate (lag_01_) and more prolonged (lag_06_) effects on cardiovascular, and respiratory mortality, respectively. That said, the onset and duration of the mechanisms of damage leading to death can vary for other health outcomes, with mortality from PM_2.5_ exposure occurring with a delay of a few days (World Health Organization. Regional Office for Europe, 2021). Little has been documented about the timing of when the largest single lag effects of PM_2.5_ on cause-specific mortality occur, and these “peaks” in mortality can be missed when reporting associations only for lag_01_, or lag_06_, and this information can provide meaningful information for health professionals. Figure 3 shows the largest single lag associations for two cause-specific mortality outcomes (hemorrhagic stroke and influenza and pneumonia); and to complement this information, results for all single lag and cumulative associations for all cause-specific mortality outcomes are reported in Table S2. Same-day exposure to PM_2.5_ (lag_0_) showed the largest single lag associations with diseases of the arteries [4.62% (95%CI: 0.73%–8.67%)], hemorrhagic stroke [2.98% (95%CI: 1.09%–4.90%)], hypertensive disease [1.34% (95%CI: 0.02%–2.68%)], and acute ischemic heart disease [0.64% (95%CI: -0.02%–1.31%)]. Lag_1_ showed the largest association with diseases of the esophagus, stomach and duodenum [3.40% (95%CI: 0.26%–6.64%)], and chronic respiratory disease [1.72% (95%CI: 0.44%–3.02%)]. Lag_4_ had the largest effects for chronic ischemic heart disease [3.74% (95%CI: 1.37%–6.16%)], and ischemic stroke [3.08% (95%CI: -0.22%–6.50%)], and lag_6_ for influenza and pneumonia [2.19% (95%CI: 0.88%–3.52%)].

**Figure 3.**
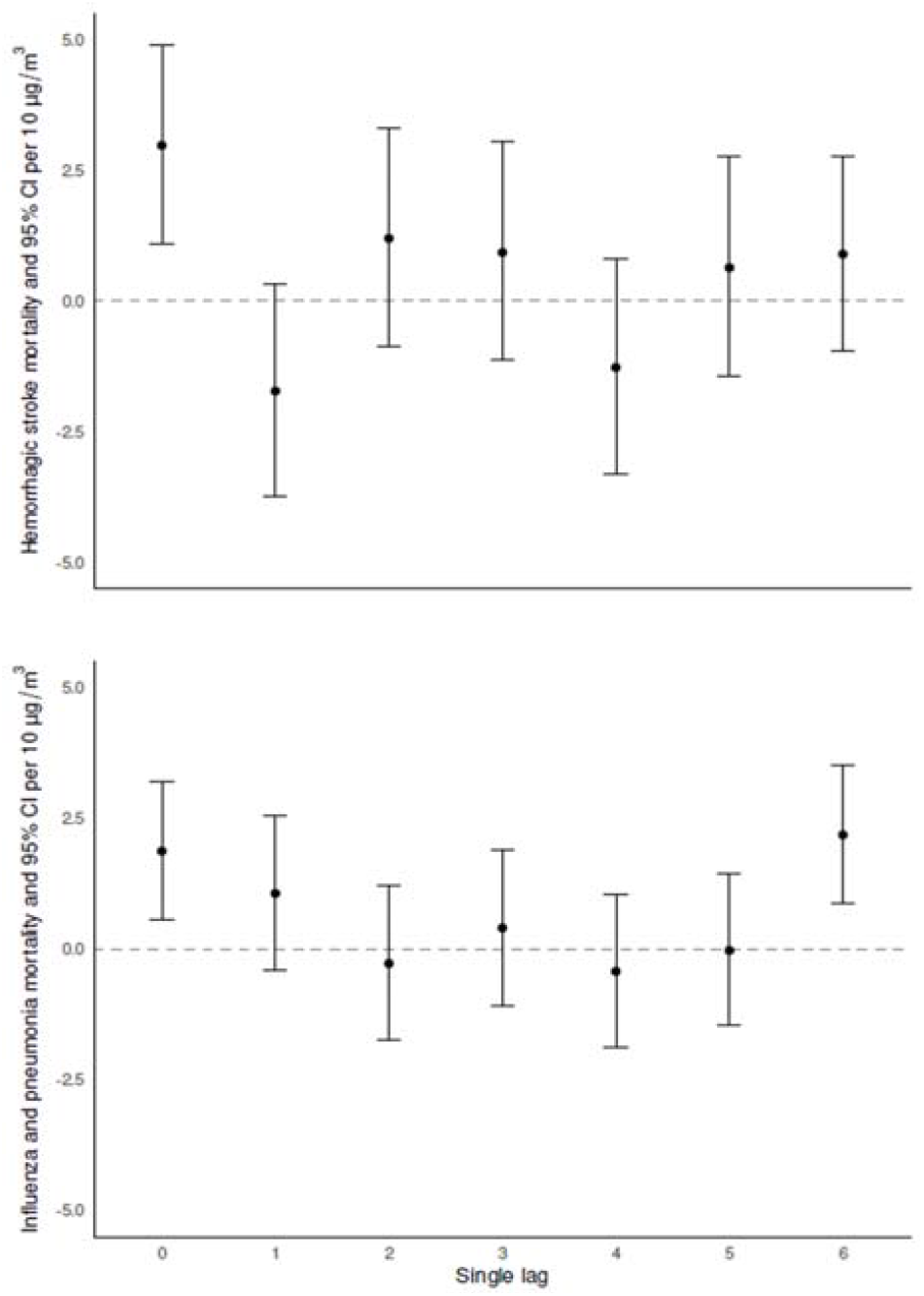
Single lag associations as percent increase (%) and 95% CI for hemorrhagic-stroke and influenza and pneumonia per 10 μg/m^3^ increase of PM_2.5_

Figure 4 shows the associations between exposure to PM_2.5_ with non-accidental mortality in adults (≥18-64 years-old) and the elderly (≥65 years-old), stratified by education level (having or not basic education), health insurance affiliation (having or not health social security affiliation), and employment status (either being working or not when the person died). Associations for lag_06_ in those with no education were 2.70% (95%CI: 0.18%–5.28%), and 2.33% (95%CI: 1.16%–3.52%) for adults and the elderly, respectively. While for those having at least basic education the associations were 1.64% (95%CI: 0.97%–2.31%) for adults, and 1.55% (95%CI: 1.00%–2.10%)], for the elderly. For the same exposure window, mortality risk in adults without health social security affiliation was 2.19% (95%CI: 1.22%–3.16%), and 1.75% (95%CI: 0.82%–2.68%) for the elderly. Adults with health insurance showed a 1.22% (95%CI: 0.35%–2.09%) mortality risk, and the elderly 1.74% (95%CI: 1.15%–2.34%). Results by employment status for the period from 2013-2019, showed that associations for adults not working at the time of death were 1.68% (95%CI: 0.36%–3.02%), and 1.83% (95%CI: 1.03%–2.63%) for the elderly; compared to those who were working with mortality risks of 0.90% (95%CI: -0.48%–2.29%) and 1.70% (95%CI: 0.23%–3.20%), for adults and the elderly, respectively. Our test of heterogeneity in effect size between SES related strata, does not suggest effect modification between SES strata. Figure S1, shows job-specific associations with PM_2.5_ exposure for the same period of time.

**Figure 4.**
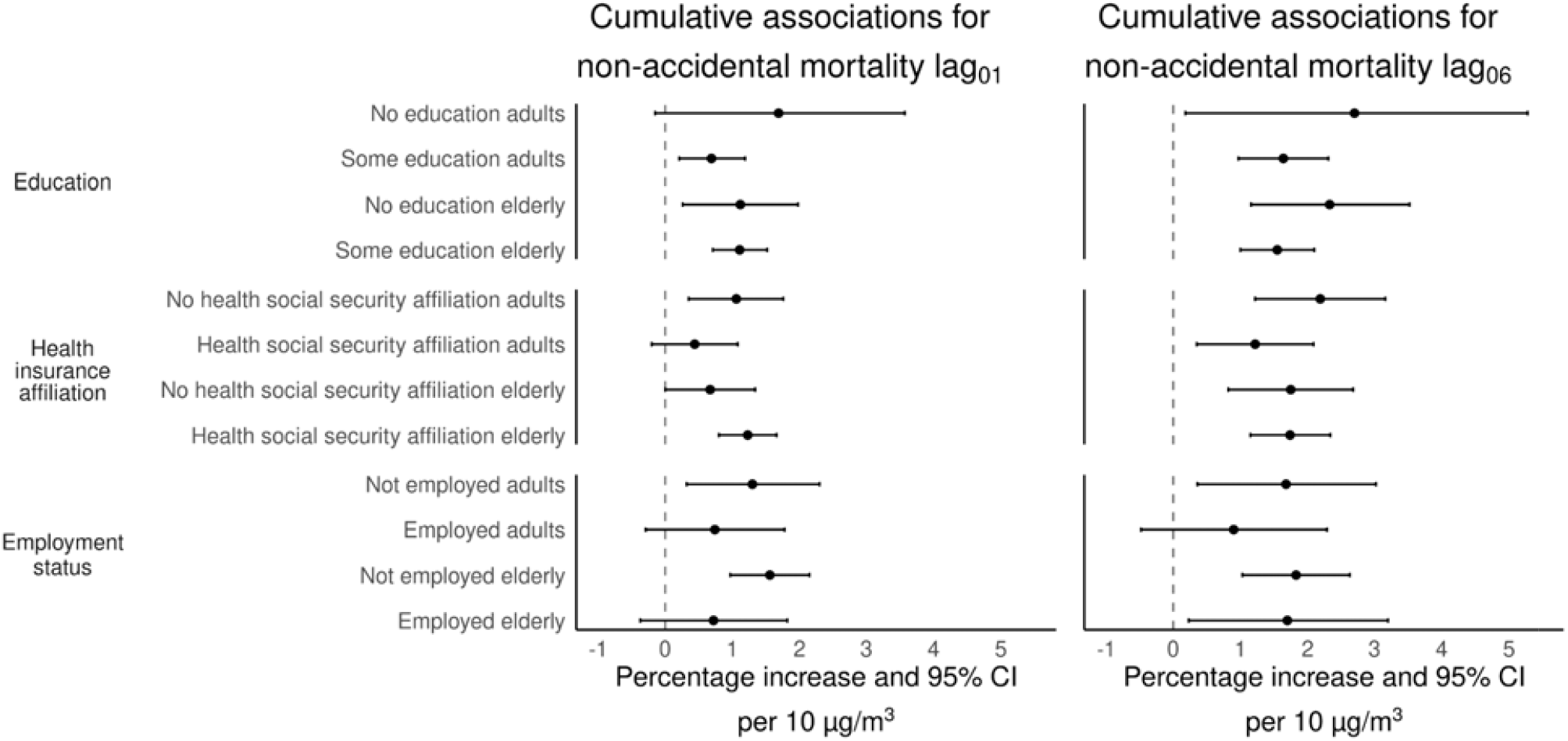
Cumulative percentage increase (%) and 95% CI for non-accidental mortality by education, health insurance affiliation, and employment status (adults 18-64 years-old, elderly ≥65 years-old) per 10 μg/m^3^ increase in PM_2.5_ over two (lag_01_) days and one week (lag_06_) in the Mexico City Metropolitan Area for 2004-2019. Associations for employment status are for the period from 2013-2019.

## Discussion

This study presents a detailed analysis on the associations between short-term exposure to PM_2.5_ with total non-external, broad-catergory (organ-system) and cause-specific mortality outcomes; and the role that age, sex, and potential SES-related effect modifiers play in such associations in the Mexico City Metropolitan Area. Our research involved the use of state-of-the-art methods to estimate daily exposure to PM_2.5_, using highly spatially-resolved satellite-based PM_2.5_ predictions. With our PM_2.5_ predictions, we were able to assign daily exposure to PM_2.5_ to all case and control days at the sub-municipality level; leveraging the smallest geographic identifier for place of residence included in the Mexican mortality records. To our knowledge, this is the most comprehensive analysis in Central Mexico in terms of geographic extent, amount of mortality outcomes analyzed, and assessment of potential effect modifiers in these associations using individual mortality records.

Our results support previous evidence on the associations between exposure to PM_2.5_ with increased risks of non-accidental, cardiovascular, cerebrovascular and respiratory mortality in the Mexico City region (Borja-Aburto et al., 1998; Gutiérrez-Avila et al., 2018). We also found associations between PM_2.5_ exposure with causes of death not previously reported in the Mexico City Metropolitan Area, such as digestive and genitourinary outcomes. Our results for non-accidental, cardiovascular and respiratory mortality in adults, the three more consistent broad-category mortality outcomes linked to PM_2.5_ exposure for lag_01_, were larger than the associations reported by Li et al. (2018) for non-accidental [0.25% (95% CI: 0.11%–0.38%)], circulatory system [0.39% (95% CI: 0.21%–0.58%)], and respiratory [0.43%; 95% CI: 0.05%–0.78%)] related mortality, and also larger than the results from Xu et al. (2020) for all-cause [0.13% (95% CI: 0.01%–0.27%)], cardiovascular [0.02% (95% CI: -0.17%–0.21%)], and respiratory [0.81% (95% CI: 0.39%–0.96%)] mortality for Beijing. Our results for the same health outcomes were similar to those reported in the meta-analyzes of Atkinson et al. (2014) for all-cause [1.04% (95% CI: 0.52%–1.56%)], cardiovascular [0.84% (95% CI: 0.41%–1.28%)], and respiratory [1.51% (95% CI: 1.01%–2.01%)] mortality (Atkinson et al., 2014). Orellano et al. (2020) also reported similar associations for all-cause [0.65% (95% CI: 0.44%–0.86%)] and cardiovascular [0.92% (95% CI: 0.61%–1.23%)] mortality, but their associations for respiratory [0.73% (95% CI: 0.29%–1.16%)] and cerebrovascular [0.72% (95% CI: 0.12%–1.32%)] mortality were lower compared to our results (Orellano et al., 2020).

However, it is possible that the selection of heterogeneous effect estimates from different lags structures that were combined in the meta-analyzes of Atkinson et al. (2014), and Orellano et al. (2020) could have induced some biases. Differential toxicity related to PM_2.5_ composition, variation in the underlying health status of the population, and distribution of potential effect modifiers between study regions can also play a role in the differences reported in the literature. Also, it has been suggested that at higher concentrations of PM_2.5_ like those observed in Asian cities, the slopes in the concentration-response function between PM_2.5_ and mortality flattens out (supralinear exposure-response curve) for cardiorespiratory outcomes, which might also explain the differences in effect estimates between study regions (Liu et al., 2019). Future research efforts exploring the shape of the concentration response functions for mortality outcomes other than cardiorespiratory outcomes could aid to understand the differences in mortality risks across study regions.

Among the cause-specific mortality associations with PM_2.5_, those related to the circulatory system (e.g. diseases of the arteries, hypertensive disease, acute ischemic heart disease, and stroke) were the most clear, with the largest associations on the same day of exposure (lag_0_ in Table S2). Li et al. (2018) also reported consistent associations between PM_2.5_ with mortality outcomes from the circulatory and respiratory systems in Beijing. However, we found larger associations compared to those reported by Li et al. (2018) for ischemic heart disease (0.46%; 95% CI: 0.19%–0.72%), influenza and pneumonia (0.35%; 95% CI: -0.32%–1.03%), and chronic respiratory disease (0.62%; 95% CI: 0.08%–1.16%). Li et al. (2018) also reported associations between PM_2.5_ and sub-specific causes of ischemic heart disease such as: acute ischemic heart disease (ICD-10 codes: I20-I22, I24), acute myocardial infarction (ICD-10 codes: I21-I22), and myocardial infarction (ICD-10 codes: I21-I23). We did not explore such subtypes of ischemic heart disease, as Li et al. (2018) did, because the grouping of such health outcomes involves the overlap of several ICD-10 codes; while we only focused on reporting associations for mutually exclusive health outcomes. We found larger associations than those reported by Xu et al. (2020) for ischemic heart disease (−0.06%; 95% CI: -0.33%–0.22%), and chronic respiratory disease (0.96%; 95% CI: 0.35%– 1.57%) also in Beijing. Our results for specific cardiovascular and cerebrovascular outcomes were larger than those reported by Chen et al. (2018) for 30 Chinese counties (Chen et al., 2018). Intentional-self harm mortality was not associated with PM_2.5_ exposure in our study, which is consistent with results of Astudillo-García et al. (2019) from a time-series study focused on suicides and exposure to different air pollutants including PM_2.5_ in Mexico City (Astudillo-García et al., 2019).

Contrary to the results from Li et al. (2018), we did not find associations between short-term exposure to PM_2.5_ with acute mortality outcomes from the nervous system (e.g. extrapyramidal and movement disorders), however we observed positive associations between cumulative exposure to PM_2.5_ with digestive (diseases of the liver) and genitourinary (renal failure) mortality. Exposure to PM_2.5_ has been associated with increased serum levels of hepatic enzymes, such as γ-glutamyltranspeptidase (GTP), and alanine transaminase (ALT), which are biomarkers of liver damage. PM_2.5_ can induce systemic oxidative stress and inflammation when deposited in the airways, and when swallowed PM_2.5_ are removed from the airways by mucociliary clearance leading to gastrointestinal exposure with the largest internal dose and adverse effects observed in the liver (Kim et al., 2014; Tavera Busso et al., 2020). On the other hand, recent evidence has shown that PM_2.5_ is related to renal injury and increases the risk of nephropathy, as well as increased risk of first hospital admission from kidney and total urinary system diseases (Lee et al., 2022; Rasking et al., 2022). PM_2.5_ can unbalance the kidney function by accumulation in the kidney tissue, endothelial dysfunction, abnormal renin-angiotensin system, and immune complex deposition. The mechanisms of damage from PM_2.5_ to the kidney involve inflammation, oxidative stress, apoptosis, DNA damage, and autophagy (Xu et al., 2022).

The most recent version of the EPA’s Integrated Science Assessment (ISA) for Particulate Matter (2019) has defined the associations between short-term exposure to PM_2.5_ with cardiovascular and respiratory mortality as causal and likely causal, respectively; but less evidence is available for making conclusions about other causes of death (EPA, 2019). Although our results suggest that short-term exposure to PM_2.5_ may trigger cause-specific mortality beyond the circulatory and respiratory systems, more epidemiological evidence is needed to understand the links between PM_2.5_ exposure with the different specific death causes evaluated in our study. Emerging evidence about the adverse effects of PM_2.5_ exposure on multiple prevalent but rarely studied causes of hospital admissions (Wei et al., 2019) adds to the body of epidemiologic evidence that supports our findings, and it opens the possibility to replicate such results in different locations with ongoing or future health studies.

Information on the timing when health effects occur after initial exposure to PM_2.5_, or any other air pollutant, is of importance for public health administrators and pollution control managers. For instance, identifying the lag days over which health effects are observed can inform the development of air quality standards, risk communication tools (e.g. air quality indices), quantification of health impacts, mitigation strategies (e.g. planning and allocation of clinical resources), and the design of emission control measures. Our results are consistent with the epidemiologic evidence showing that short-term exposure to PM_2.5_ has an immediate effect on non-accidental and cardiovascular mortality, and a more prolonged effect on respiratory mortality (EPA, 2019). However, when considering specific death causes (Table S2) within the broad-category mortality outcomes, there seems to be more variability about the timing when the largest (single and cumulative) associations are observed. For instance, chronic ischemic heart disease and ischemic stroke showed the largest associations with PM_2.5_ on lag_4_, while the rest of the death causes from the circulatory system showed the largest association on lag_0_ and lag_1_. For respiratory mortality, our results are within the range of observed associations, with chronic respiratory disease showing the largest association with PM_2.5_ on lag_1_, while the association with influenza and pneumonia remained high at lag_6_. On the other hand, it is known that the strength of the relationship between exposure to PM_2.5_ and health effects varies depending on the exposure duration i.e., short- or long-term exposure, but less is known about the strength of this relationship for sub-daily exposures e.g. hourly peak exposures. Few epidemiologic studies have focused on the associations between sub-daily exposures to PM_2.5_ with morbidity and mortality outcomes (Lin et al., 2017; Madsen et al., 2012). Results from such investigations are inconclusive on whether sub-daily exposures pose a greater risk of mortality, or not, compared to daily averaged exposures; being possible that the sub-daily metrics of PM_2.5_ employed for exposure assessment in previous studies had a limited spatiotemporal variability (EPA, 2019). In our research, we only focused on exposures to daily mean PM_2.5_ concentrations assuming that the same averaging time could have the same relevance for all mortality outcomes. However, we can not rule out the possibility that there might be averaging times (sub-daily, or sub-chronic exposures) that could be of greater relevance for the different death causes analyzed in our research.

It is known that subgroups of the population with lower SES indicators may have higher prevalence of health disorders and comorbidities, lower living standards, and higher exposure to air pollutants (e.g. living closer to a highway or lack of green spaces) (Shavers, 2007; Xu et al., 2020). We used three indicators of SES, namely education, health social security affiliation, and employment status to assess their roles as potential effect modifiers in the PM_2.5_-mortality relationship. Although in general point estimates for the lower SES related strata (lower education level, not having health social security affiliation, and not being working at the time of death) seemed larger than those in the higher SES indicators; we did not find evidence of effect modification in the PM_2.5_-mortality associations (analyzed on the log scale) between SES strata (Altman and Bland, 2003). In Mexico there is limited evidence about the role of education in the association between short-term exposure to PM_2.5_ and mortality, however O’Neill et al. (2008) did not observe evidence of heterogeneity in the association between short-term exposure to PM_10_ and mortality across education levels (O’Neill et al., 2008). Health insurance access can modify the susceptibility to detrimental effects of environmental stressors, including PM_2.5_, by providing access to medications, supplements and overall preventive services (Woolhandler and Himmelstein, 2017). Our results did not show significant effect modification between strata of health social security affiliation. Also, our results for employment status did not suggest greater risk of mortality for those in the not-employed strata compared with people in the employed category. The evidence on the effects of working status on health suggests that unemployment is related to poor health (e.g. greater risk of mental illness, physical complaints, and an increased risk for coronary heart diseases) and early mortality (Kim and von dem Knesebeck, 2015). Although our models were further stratified by age group to reduce its potential confounding effects, our results should be carefully interpreted, since there was not a well-defined measure of unemployment excluding all the economically inactive (Clemens et al., 2015). Also, we did not have information on the health status of those in the “non-working” group before death, so it is not possible to determine if a deteriorating health status led to a “non-working” condition (healthy worker effect). When evaluating the role of specific job types in the association between short-term exposure to PM_2.5_ and mortality, the lack of consistency in the codification of specific job categories throughout the period of study led us to evaluate these associations for a shorter time period (2013-2019), likely reducing power to identify associations by individual job categories (Figure S1). Two major problems with the use of death certificates for classifying employment include ambiguity about whether unemployed persons are actually retired or if a prior occupation is reported for retired decedents and thus may not represent their recent exposures.

Other limitations in our study could be related to the inaccurate classification of death causes for some diseases. In general, inaccurate classification of death causes in Mexico is considered low (Naghavi et al., 2010). However, errors might exist in the classification of some mortality outcomes, including those from the circulatory system, which have been recognized among the most affected by the use of “garbage codes”, i.e. those that are not underlying causes of death such as heart failure, but codes that define poorly-specified diagnoses not clearly identifying a death cause. For Mexico City, we have previously reported that the use of garbage codes tends to bias effect estimates of cardiovascular outcomes to the null, and that proportional redistribution of garbage codes into actual death causes may help to reduce such bias (Gutiérrez-Avila et al., 2018). Also, there are contributions to exposure measurement error that accumulate from the combination of prediction model error and the use of locality of residence to represent all relevant time-activity, which may bias effect estimates (albeit less than the use of a single city-wide exposure time series). Finally, the study of effect modification by cause-specific mortality, age group, sex, and SES indicators can be sensitive to analytic methods. Although our stratified analyses have been the standard in case-crossover and time-series studies, the sparse numbers in some of the strata could have reduced statistical power to detect associations.

## Conclusion

Our findings support the growing evidence that short-term exposure to PM_2.5_ may trigger cause-specific mortality beyond cardiovascular and respiratory outcomes, including mortality from diseases of the digestive and genitourinary systems. This evidence can be used to inform and update local environmental health policies, and to more comprehensively assess the benefits of interventions aimed to reduce PM_2.5_ pollution in the Mexico City Metropolitan Area. Our findings suggest that short-term exposure to PM_2.5_ increases the risk of acute mortality in the Mexico City Metropolitan Area, regardless of age groups, and SES characteristics.

## Data Availability

All data used in the present study are available upon reasonable request to the authors

## Ethical Approval

Because the records used in all analyses were already publicly available, the senior author (ACJ) received a determination of exempt human research [45 CFR 46. 101(b) (Category 4)] from the Institutional Review Board of the Icahn School of Medicine at Mount Sinai.

