## Supplemental material for "Daily exposure to PM_2.5_ and 1.5 million deaths: A time-stratified case-crossover analysis in the Mexico City Metropolitan Area"

Figure S1. Cumulative percentage increase (%) and 95% CI for non-accidental mortality by education, insurance type and job categories (adults ≥18 years-old) per 10 μg/m^3^ increase in PM_2.5_ over two (lag_01_) days and one week (lag_06_) in the Mexico City Metropolitan Area for 2004-2019. Job category associations are for the period from 2013-2019.

Table S1. Single lag and cumulative Odds Ratios and 95% confidence intervals for broad-group mortality outcomes associated with 10μg/m^3^ increase in PM_2.5_

Table S2. Single lag and cumulative Odds Ratios and 95% confidence intervals for cause-specific mortality outcomes associated with 10μg/m^3^ increase in PM_2.5_

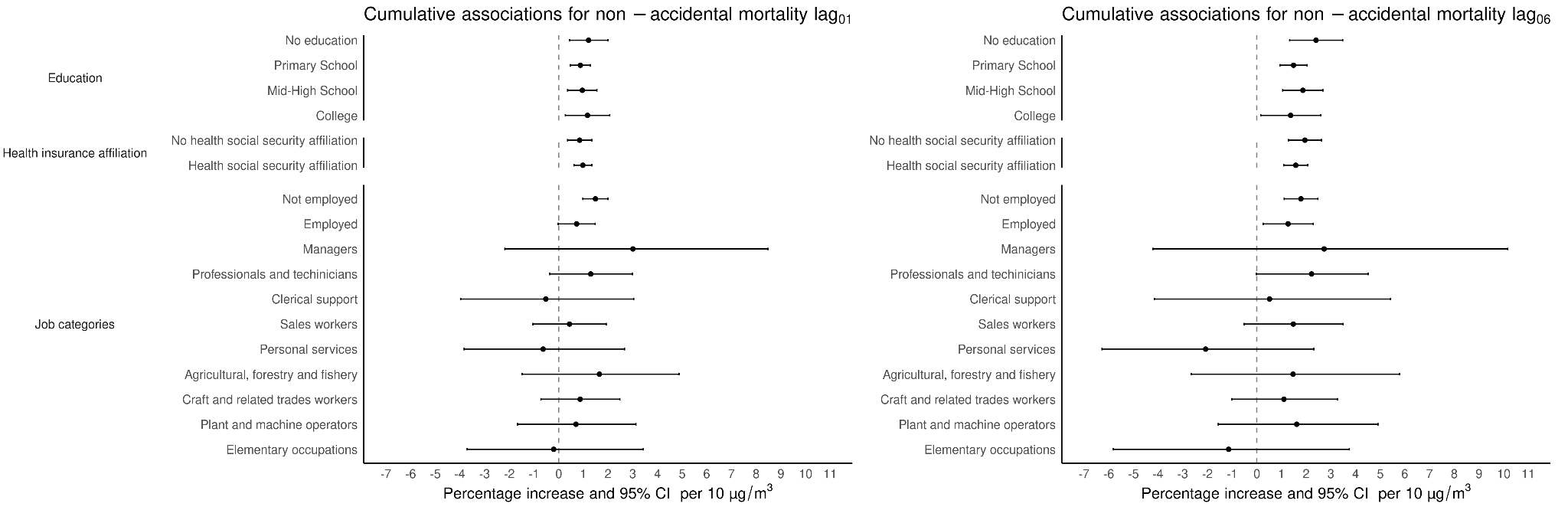

Figure S1. Cumulative percentage increase (%) and 95% CI for non-accidental mortality by education, health insurance affiliation, and job categories (adults ≥18 years-old) per 10 μg/m^3^ increase in PM_2.5_ over two (lag_01_) days and one week (lag_06_) in the Mexico City Metropolitan Area for 2004-2019. Job category associations are for the period from 2013-2019.

Table S1. Single lag and cumulative Odds Ratios and 95% confidence intervals for broad-category mortality outcomes associated with 10μg/m^3^ increase in PM_2.5_

| **Group** | **lag** | **OR** | **Lower Limit** | **Upper Limit** | **OR cumulative** | **Lower Limit cumulative** | **Upper Limit cumulative** |
| --- | --- | --- | --- | --- | --- | --- | --- |
| All causes adults (18-64 years-old) | 0 | 1.008 | 1.004 | 1.012 | 1.008 | 1.004 | 1.012 |
| All causes adults (18-64 years-old) | 1 | 0.999 | 0.994 | 1.004 | 1.007 | 1.003 | 1.012 |
| All causes adults (18-64 years-old) | 2 | 1.004 | 0.999 | 1.008 | 1.011 | 1.006 | 1.016 |
| All causes adults (18-64 years-old) | 3 | 1.002 | 0.997 | 1.006 | 1.012 | 1.007 | 1.018 |
| All causes adults (18-64 years-old) | 4 | 0.998 | 0.993 | 1.002 | 1.010 | 1.004 | 1.016 |
| All causes adults (18-64 years-old) | 5 | 1.002 | 0.997 | 1.006 | 1.012 | 1.005 | 1.018 |
| All causes adults (18-64 years-old) | 6 | 1.005 | 1.001 | 1.009 | 1.017 | 1.010 | 1.023 |
| All causes elderly (+65 years-old) | 0 | 1.005 | 1.002 | 1.008 | 1.005 | 1.002 | 1.008 |
| All causes elderly (+65 years-old) | 1 | 1.005 | 1.002 | 1.009 | 1.010 | 1.007 | 1.014 |
| All causes elderly (+65 years-old) | 2 | 0.997 | 0.993 | 1.001 | 1.007 | 1.003 | 1.011 |
| All causes elderly (+65 years-old) | 3 | 1.001 | 0.997 | 1.004 | 1.008 | 1.004 | 1.012 |
| All causes elderly (+65 years-old) | 4 | 1.003 | 0.999 | 1.006 | 1.011 | 1.006 | 1.015 |
| All causes elderly (+65 years-old) | 5 | 1.002 | 0.998 | 1.005 | 1.013 | 1.008 | 1.018 |
| All causes elderly (+65 years-old) | 6 | 1.004 | 1.001 | 1.007 | 1.017 | 1.012 | 1.022 |
| Men | 0 | 1.007 | 1.003 | 1.010 | 1.007 | 1.003 | 1.010 |
| Men | 1 | 1.002 | 0.998 | 1.006 | 1.008 | 1.004 | 1.012 |
| Men | 2 | 1.000 | 0.996 | 1.004 | 1.008 | 1.004 | 1.012 |
| Men | 3 | 1.000 | 0.996 | 1.004 | 1.008 | 1.003 | 1.012 |
| Men | 4 | 1.000 | 0.996 | 1.004 | 1.008 | 1.003 | 1.013 |
| Men | 5 | 1.004 | 1.000 | 1.008 | 1.012 | 1.006 | 1.017 |
| Men | 6 | 1.006 | 1.002 | 1.009 | 1.017 | 1.012 | 1.023 |
| Women | 0 | 1.006 | 1.002 | 1.010 | 1.006 | 1.002 | 1.010 |
| Women | 1 | 1.004 | 1.000 | 1.008 | 1.010 | 1.006 | 1.014 |
| Women | 2 | 0.999 | 0.995 | 1.003 | 1.009 | 1.005 | 1.014 |
| Women | 3 | 1.002 | 0.998 | 1.006 | 1.012 | 1.007 | 1.016 |
| Women | 4 | 1.002 | 0.998 | 1.006 | 1.014 | 1.008 | 1.019 |
| Women | 5 | 0.999 | 0.995 | 1.003 | 1.013 | 1.007 | 1.018 |
| Women | 6 | 1.003 | 0.999 | 1.007 | 1.016 | 1.010 | 1.021 |
| All Mental and behavioral disorders adults | 0 | 0.983 | 0.942 | 1.026 | 0.983 | 0.942 | 1.026 |
| All Mental and behavioral disorders adults | 1 | 0.998 | 0.952 | 1.047 | 0.981 | 0.935 | 1.029 |
| All Mental and behavioral disorders adults | 2 | 0.982 | 0.936 | 1.029 | 0.963 | 0.914 | 1.014 |
| All Mental and behavioral disorders adults | 3 | 0.955 | 0.910 | 1.003 | 0.920 | 0.870 | 0.973 |
| All Mental and behavioral disorders adults | 4 | 1.022 | 0.976 | 1.071 | 0.940 | 0.886 | 0.998 |
| All Mental and behavioral disorders adults | 5 | 0.995 | 0.950 | 1.043 | 0.936 | 0.879 | 0.997 |
| All Mental and behavioral disorders adults | 6 | 1.015 | 0.975 | 1.057 | 0.950 | 0.891 | 1.013 |
| All Mental and behavioral disorders elderly | 0 | 1.038 | 0.990 | 1.089 | 1.038 | 0.990 | 1.089 |
| All Mental and behavioral disorders elderly | 1 | 0.997 | 0.946 | 1.051 | 1.035 | 0.982 | 1.092 |
| All Mental and behavioral disorders elderly | 2 | 1.016 | 0.962 | 1.073 | 1.052 | 0.991 | 1.115 |
| All Mental and behavioral disorders elderly | 3 | 1.023 | 0.970 | 1.079 | 1.076 | 1.010 | 1.146 |
| All Mental and behavioral disorders elderly | 4 | 1.008 | 0.955 | 1.063 | 1.084 | 1.013 | 1.160 |
| All Mental and behavioral disorders elderly | 5 | 0.966 | 0.914 | 1.021 | 1.047 | 0.975 | 1.125 |
| All Mental and behavioral disorders elderly | 6 | 0.989 | 0.942 | 1.039 | 1.036 | 0.963 | 1.114 |
| All nervous adults | 0 | 1.029 | 1.001 | 1.058 | 1.029 | 1.001 | 1.058 |
| All nervous adults | 1 | 0.978 | 0.948 | 1.009 | 1.007 | 0.976 | 1.039 |
| All nervous adults | 2 | 1.013 | 0.981 | 1.046 | 1.020 | 0.985 | 1.056 |
| All nervous adults | 3 | 0.975 | 0.945 | 1.007 | 0.995 | 0.958 | 1.032 |
| All nervous adults | 4 | 0.983 | 0.953 | 1.014 | 0.977 | 0.940 | 1.016 |
| All nervous adults | 5 | 1.001 | 0.971 | 1.033 | 0.979 | 0.939 | 1.020 |
| All nervous adults | 6 | 0.998 | 0.971 | 1.027 | 0.977 | 0.937 | 1.019 |
| All nervous elderly | 0 | 1.002 | 0.973 | 1.032 | 1.002 | 0.973 | 1.032 |
| All nervous elderly | 1 | 1.022 | 0.988 | 1.056 | 1.023 | 0.990 | 1.058 |
| All nervous elderly | 2 | 0.993 | 0.960 | 1.026 | 1.016 | 0.979 | 1.053 |
| All nervous elderly | 3 | 0.980 | 0.948 | 1.014 | 0.996 | 0.957 | 1.035 |
| All nervous elderly | 4 | 1.023 | 0.989 | 1.058 | 1.018 | 0.977 | 1.061 |
| All nervous elderly | 5 | 0.992 | 0.959 | 1.025 | 1.010 | 0.967 | 1.055 |
| All nervous elderly | 6 | 0.977 | 0.949 | 1.006 | 0.987 | 0.944 | 1.032 |
| All cardiovascular adults | 0 | 1.020 | 1.010 | 1.030 | 1.020 | 1.010 | 1.030 |
| All cardiovascular adults | 1 | 0.990 | 0.980 | 1.001 | 1.010 | 1.000 | 1.021 |
| All cardiovascular adults | 2 | 1.004 | 0.993 | 1.014 | 1.014 | 1.002 | 1.026 |
| All cardiovascular adults | 3 | 0.996 | 0.985 | 1.006 | 1.009 | 0.997 | 1.022 |
| All cardiovascular adults | 4 | 0.996 | 0.985 | 1.006 | 1.005 | 0.991 | 1.018 |
| All cardiovascular adults | 5 | 1.000 | 0.989 | 1.010 | 1.004 | 0.991 | 1.019 |
| All cardiovascular adults | 6 | 1.009 | 1.000 | 1.019 | 1.014 | 1.000 | 1.028 |
| All cardiovascular elderly | 0 | 1.004 | 0.999 | 1.010 | 1.004 | 0.999 | 1.010 |
| All cardiovascular elderly | 1 | 1.006 | 1.000 | 1.012 | 1.010 | 1.004 | 1.016 |
| All cardiovascular elderly | 2 | 0.998 | 0.992 | 1.004 | 1.008 | 1.002 | 1.015 |
| All cardiovascular elderly | 3 | 1.000 | 0.994 | 1.006 | 1.008 | 1.001 | 1.015 |
| All cardiovascular elderly | 4 | 1.007 | 1.001 | 1.013 | 1.015 | 1.007 | 1.023 |
| All cardiovascular elderly | 5 | 1.000 | 0.994 | 1.006 | 1.015 | 1.007 | 1.024 |
| All cardiovascular elderly | 6 | 1.003 | 0.998 | 1.008 | 1.018 | 1.010 | 1.027 |
| All cerebrovascular adults | 0 | 1.032 | 1.012 | 1.053 | 1.032 | 1.012 | 1.053 |
| All cerebrovascular adults | 1 | 0.996 | 0.974 | 1.019 | 1.028 | 1.005 | 1.052 |
| All cerebrovascular adults | 2 | 0.980 | 0.958 | 1.003 | 1.008 | 0.983 | 1.034 |
| All cerebrovascular adults | 3 | 1.013 | 0.991 | 1.036 | 1.022 | 0.994 | 1.050 |
| All cerebrovascular adults | 4 | 0.997 | 0.974 | 1.020 | 1.019 | 0.990 | 1.048 |
| All cerebrovascular adults | 5 | 1.004 | 0.981 | 1.027 | 1.022 | 0.992 | 1.054 |
| All cerebrovascular adults | 6 | 1.014 | 0.993 | 1.035 | 1.036 | 1.005 | 1.069 |
| All cerebrovascular elderly | 0 | 0.994 | 0.982 | 1.006 | 0.994 | 0.982 | 1.006 |
| All cerebrovascular elderly | 1 | 1.003 | 0.990 | 1.017 | 0.997 | 0.984 | 1.011 |
| All cerebrovascular elderly | 2 | 1.000 | 0.987 | 1.014 | 0.998 | 0.983 | 1.013 |
| All cerebrovascular elderly | 3 | 0.999 | 0.985 | 1.012 | 0.996 | 0.981 | 1.012 |
| All cerebrovascular elderly | 4 | 0.999 | 0.986 | 1.013 | 0.996 | 0.979 | 1.013 |
| All cerebrovascular elderly | 5 | 1.006 | 0.992 | 1.019 | 1.001 | 0.983 | 1.019 |
| All cerebrovascular elderly | 6 | 1.005 | 0.993 | 1.017 | 1.006 | 0.988 | 1.024 |
| All respiratory adults | 0 | 1.009 | 0.992 | 1.025 | 1.009 | 0.992 | 1.025 |
| All respiratory adults | 1 | 1.016 | 0.998 | 1.035 | 1.025 | 1.006 | 1.044 |
| All respiratory adults | 2 | 0.988 | 0.970 | 1.006 | 1.013 | 0.992 | 1.034 |
| All respiratory adults | 3 | 1.005 | 0.987 | 1.024 | 1.018 | 0.996 | 1.041 |
| All respiratory adults | 4 | 1.003 | 0.985 | 1.021 | 1.021 | 0.997 | 1.045 |
| All respiratory adults | 5 | 0.993 | 0.975 | 1.012 | 1.014 | 0.989 | 1.039 |
| All respiratory adults | 6 | 1.012 | 0.995 | 1.029 | 1.026 | 1.000 | 1.052 |
| All respiratory elderly | 0 | 1.002 | 0.993 | 1.011 | 1.002 | 0.993 | 1.011 |
| All respiratory elderly | 1 | 1.018 | 1.008 | 1.028 | 1.020 | 1.010 | 1.030 |
| All respiratory elderly | 2 | 0.993 | 0.983 | 1.003 | 1.013 | 1.002 | 1.024 |
| All respiratory elderly | 3 | 1.004 | 0.994 | 1.014 | 1.017 | 1.005 | 1.029 |
| All respiratory elderly | 4 | 0.996 | 0.986 | 1.006 | 1.013 | 1.000 | 1.026 |
| All respiratory elderly | 5 | 1.002 | 0.992 | 1.012 | 1.015 | 1.001 | 1.029 |
| All respiratory elderly | 6 | 1.019 | 1.010 | 1.028 | 1.035 | 1.020 | 1.049 |
| All digestive adults | 0 | 1.008 | 0.998 | 1.019 | 1.008 | 0.998 | 1.019 |
| All digestive adults | 1 | 0.997 | 0.985 | 1.009 | 1.006 | 0.994 | 1.017 |
| All digestive adults | 2 | 1.016 | 1.004 | 1.028 | 1.022 | 1.008 | 1.035 |
| All digestive adults | 3 | 0.998 | 0.986 | 1.010 | 1.019 | 1.005 | 1.034 |
| All digestive adults | 4 | 0.999 | 0.987 | 1.011 | 1.018 | 1.003 | 1.034 |
| All digestive adults | 5 | 1.006 | 0.994 | 1.018 | 1.024 | 1.009 | 1.041 |
| All digestive adults | 6 | 0.996 | 0.986 | 1.007 | 1.021 | 1.005 | 1.037 |
| All digestive elderly | 0 | 1.004 | 0.994 | 1.015 | 1.004 | 0.994 | 1.015 |
| All digestive elderly | 1 | 1.008 | 0.996 | 1.020 | 1.012 | 1.000 | 1.024 |
| All digestive elderly | 2 | 0.994 | 0.982 | 1.006 | 1.005 | 0.992 | 1.018 |
| All digestive elderly | 3 | 0.996 | 0.984 | 1.008 | 1.001 | 0.988 | 1.015 |
| All digestive elderly | 4 | 1.004 | 0.992 | 1.016 | 1.005 | 0.991 | 1.020 |
| All digestive elderly | 5 | 0.994 | 0.983 | 1.006 | 1.000 | 0.984 | 1.015 |
| All digestive elderly | 6 | 1.008 | 0.998 | 1.019 | 1.008 | 0.992 | 1.024 |
| All genitourinary adults | 0 | 0.996 | 0.975 | 1.017 | 0.996 | 0.975 | 1.017 |
| All genitourinary adults | 1 | 0.996 | 0.972 | 1.020 | 0.992 | 0.969 | 1.015 |
| All genitourinary adults | 2 | 0.985 | 0.962 | 1.009 | 0.977 | 0.952 | 1.003 |
| All genitourinary adults | 3 | 1.047 | 1.023 | 1.072 | 1.023 | 0.995 | 1.052 |
| All genitourinary adults | 4 | 1.001 | 0.978 | 1.025 | 1.024 | 0.994 | 1.054 |
| All genitourinary adults | 5 | 0.996 | 0.972 | 1.020 | 1.019 | 0.988 | 1.052 |
| All genitourinary adults | 6 | 1.014 | 0.993 | 1.036 | 1.034 | 1.001 | 1.067 |
| All genitourinary elderly | 0 | 1.005 | 0.988 | 1.021 | 1.005 | 0.988 | 1.021 |
| All genitourinary elderly | 1 | 1.007 | 0.988 | 1.025 | 1.011 | 0.993 | 1.030 |
| All genitourinary elderly | 2 | 1.004 | 0.985 | 1.022 | 1.015 | 0.995 | 1.036 |
| All genitourinary elderly | 3 | 0.999 | 0.981 | 1.018 | 1.014 | 0.992 | 1.036 |
| All genitourinary elderly | 4 | 0.998 | 0.980 | 1.017 | 1.012 | 0.990 | 1.036 |
| All genitourinary elderly | 5 | 1.011 | 0.993 | 1.030 | 1.024 | 0.999 | 1.048 |
| All genitourinary elderly | 6 | 0.999 | 0.983 | 1.016 | 1.023 | 0.998 | 1.048 |

Table S2. Single lag and cumulative Odds Ratios and 95% confidence intervals for cause-specific mortality outcomes associated with 10μg/m^3^ increase in PM_2.5_

| **Group** | **lag** | **OR** | **Lower Limit** | **Upper Limit** | **OR cumulative** | **Lower Limit cumulative** | **Upper Limit cumulative** |
| --- | --- | --- | --- | --- | --- | --- | --- |
| Extrapyramidal and movement disorders | 0 | 0.991 | 0.942 | 1.043 | 0.991 | 0.942 | 1.043 |
| Extrapyramidal and movement disorders | 1 | 1.048 | 0.988 | 1.111 | 1.039 | 0.980 | 1.101 |
| Extrapyramidal and movement disorders | 2 | 0.966 | 0.910 | 1.025 | 1.003 | 0.941 | 1.070 |
| Extrapyramidal and movement disorders | 3 | 0.991 | 0.936 | 1.049 | 0.994 | 0.928 | 1.064 |
| Extrapyramidal and movement disorders | 4 | 1.027 | 0.970 | 1.087 | 1.020 | 0.949 | 1.096 |
| Extrapyramidal and movement disorders | 5 | 0.976 | 0.922 | 1.034 | 0.996 | 0.922 | 1.075 |
| Extrapyramidal and movement disorders | 6 | 1.000 | 0.950 | 1.053 | 0.996 | 0.921 | 1.076 |
| Chronic rheumatic heart disease | 0 | 0.978 | 0.932 | 1.026 | 0.978 | 0.932 | 1.026 |
| Chronic rheumatic heart disease | 1 | 1.035 | 0.982 | 1.091 | 1.012 | 0.960 | 1.067 |
| Chronic rheumatic heart disease | 2 | 1.013 | 0.964 | 1.065 | 1.025 | 0.968 | 1.085 |
| Chronic rheumatic heart disease | 3 | 1.014 | 0.964 | 1.067 | 1.039 | 0.976 | 1.106 |
| Chronic rheumatic heart disease | 4 | 1.021 | 0.971 | 1.075 | 1.061 | 0.993 | 1.135 |
| Chronic rheumatic heart disease | 5 | 0.999 | 0.950 | 1.051 | 1.060 | 0.989 | 1.138 |
| Chronic rheumatic heart disease | 6 | 0.948 | 0.905 | 0.992 | 1.005 | 0.935 | 1.080 |
| Hypertensive diseases | 0 | 1.013 | 1.000 | 1.027 | 1.013 | 1.000 | 1.027 |
| Hypertensive diseases | 1 | 0.998 | 0.984 | 1.013 | 1.011 | 0.997 | 1.026 |
| Hypertensive diseases | 2 | 1.002 | 0.987 | 1.016 | 1.013 | 0.997 | 1.029 |
| Hypertensive diseases | 3 | 1.006 | 0.992 | 1.021 | 1.019 | 1.002 | 1.037 |
| Hypertensive diseases | 4 | 0.997 | 0.982 | 1.012 | 1.016 | 0.998 | 1.035 |
| Hypertensive diseases | 5 | 1.001 | 0.987 | 1.016 | 1.018 | 0.998 | 1.038 |
| Hypertensive diseases | 6 | 1.005 | 0.992 | 1.018 | 1.023 | 1.003 | 1.043 |
| Acute ischemic heart disease | 0 | 1.006 | 1.000 | 1.013 | 1.006 | 1.000 | 1.013 |
| Acute ischemic heart disease | 1 | 1.006 | 0.998 | 1.013 | 1.012 | 1.005 | 1.020 |
| Acute ischemic heart disease | 2 | 0.999 | 0.992 | 1.007 | 1.011 | 1.003 | 1.020 |
| Acute ischemic heart disease | 3 | 0.997 | 0.989 | 1.004 | 1.008 | 0.999 | 1.017 |
| Acute ischemic heart disease | 4 | 1.005 | 0.997 | 1.012 | 1.013 | 1.004 | 1.023 |
| Acute ischemic heart disease | 5 | 0.999 | 0.992 | 1.006 | 1.012 | 1.002 | 1.022 |
| Acute ischemic heart disease | 6 | 1.004 | 0.998 | 1.011 | 1.016 | 1.006 | 1.026 |
| Chronic ischemic heart disease | 0 | 1.023 | 1.002 | 1.044 | 1.023 | 1.002 | 1.044 |
| Chronic ischemic heart disease | 1 | 0.992 | 0.969 | 1.016 | 1.015 | 0.992 | 1.039 |
| Chronic ischemic heart disease | 2 | 1.012 | 0.988 | 1.036 | 1.027 | 1.000 | 1.054 |
| Chronic ischemic heart disease | 3 | 0.973 | 0.950 | 0.996 | 0.999 | 0.971 | 1.027 |
| Chronic ischemic heart disease | 4 | 1.037 | 1.014 | 1.062 | 1.036 | 1.006 | 1.067 |
| Chronic ischemic heart disease | 5 | 0.984 | 0.962 | 1.007 | 1.020 | 0.988 | 1.052 |
| Chronic ischemic heart disease | 6 | 1.004 | 0.984 | 1.025 | 1.024 | 0.992 | 1.058 |
| Pulmonary heart disease | 0 | 1.021 | 0.985 | 1.059 | 1.021 | 0.985 | 1.059 |
| Pulmonary heart disease | 1 | 0.998 | 0.958 | 1.040 | 1.019 | 0.978 | 1.062 |
| Pulmonary heart disease | 2 | 0.998 | 0.958 | 1.040 | 1.018 | 0.972 | 1.065 |
| Pulmonary heart disease | 3 | 1.004 | 0.964 | 1.045 | 1.021 | 0.972 | 1.073 |
| Pulmonary heart disease | 4 | 0.994 | 0.953 | 1.037 | 1.015 | 0.963 | 1.070 |
| Pulmonary heart disease | 5 | 1.000 | 0.960 | 1.042 | 1.015 | 0.960 | 1.073 |
| Pulmonary heart disease | 6 | 0.999 | 0.963 | 1.036 | 1.014 | 0.958 | 1.072 |
| Other forms of heart disease | 0 | 1.006 | 0.988 | 1.024 | 1.006 | 0.988 | 1.024 |
| Other forms of heart disease | 1 | 0.999 | 0.979 | 1.019 | 1.005 | 0.985 | 1.025 |
| Other forms of heart disease | 2 | 1.004 | 0.984 | 1.024 | 1.008 | 0.987 | 1.031 |
| Other forms of heart disease | 3 | 1.004 | 0.985 | 1.024 | 1.013 | 0.989 | 1.037 |
| Other forms of heart disease | 4 | 0.997 | 0.978 | 1.017 | 1.010 | 0.985 | 1.035 |
| Other forms of heart disease | 5 | 1.011 | 0.991 | 1.031 | 1.021 | 0.994 | 1.048 |
| Other forms of heart disease | 6 | 1.003 | 0.985 | 1.021 | 1.024 | 0.997 | 1.052 |
| Stroke hemorrhagic | 0 | 1.030 | 1.011 | 1.049 | 1.030 | 1.011 | 1.049 |
| Stroke hemorrhagic | 1 | 0.983 | 0.963 | 1.003 | 1.012 | 0.992 | 1.033 |
| Stroke hemorrhagic | 2 | 1.012 | 0.991 | 1.033 | 1.024 | 1.001 | 1.048 |
| Stroke hemorrhagic | 3 | 1.009 | 0.989 | 1.030 | 1.034 | 1.009 | 1.059 |
| Stroke hemorrhagic | 4 | 0.987 | 0.967 | 1.008 | 1.021 | 0.994 | 1.047 |
| Stroke hemorrhagic | 5 | 1.006 | 0.986 | 1.028 | 1.027 | 0.999 | 1.056 |
| Stroke hemorrhagic | 6 | 1.009 | 0.991 | 1.028 | 1.036 | 1.008 | 1.065 |
| Stroke ischemic | 0 | 1.005 | 0.976 | 1.035 | 1.005 | 0.976 | 1.035 |
| Stroke ischemic | 1 | 1.021 | 0.988 | 1.055 | 1.026 | 0.993 | 1.060 |
| Stroke ischemic | 2 | 1.005 | 0.972 | 1.039 | 1.031 | 0.994 | 1.069 |
| Stroke ischemic | 3 | 0.992 | 0.960 | 1.026 | 1.023 | 0.984 | 1.063 |
| Stroke ischemic | 4 | 1.031 | 0.998 | 1.065 | 1.055 | 1.012 | 1.099 |
| Stroke ischemic | 5 | 0.984 | 0.952 | 1.017 | 1.038 | 0.993 | 1.084 |
| Stroke ischemic | 6 | 0.999 | 0.970 | 1.029 | 1.037 | 0.992 | 1.083 |
| Diseases of arteries | 0 | 1.046 | 1.007 | 1.087 | 1.046 | 1.007 | 1.087 |
| Diseases of arteries | 1 | 0.973 | 0.931 | 1.017 | 1.018 | 0.975 | 1.063 |
| Diseases of arteries | 2 | 0.960 | 0.919 | 1.003 | 0.978 | 0.932 | 1.025 |
| Diseases of arteries | 3 | 1.001 | 0.959 | 1.044 | 0.978 | 0.930 | 1.030 |
| Diseases of arteries | 4 | 1.033 | 0.990 | 1.077 | 1.010 | 0.957 | 1.067 |
| Diseases of arteries | 5 | 0.944 | 0.903 | 0.987 | 0.954 | 0.900 | 1.012 |
| Diseases of arteries | 6 | 1.032 | 0.991 | 1.074 | 0.984 | 0.928 | 1.044 |
| Influenza and pneumonia | 0 | 1.019 | 1.006 | 1.032 | 1.019 | 1.006 | 1.032 |
| Influenza and pneumonia | 1 | 1.011 | 0.996 | 1.026 | 1.030 | 1.015 | 1.045 |
| Influenza and pneumonia | 2 | 0.997 | 0.983 | 1.012 | 1.027 | 1.010 | 1.044 |
| Influenza and pneumonia | 3 | 1.004 | 0.989 | 1.019 | 1.031 | 1.013 | 1.049 |
| Influenza and pneumonia | 4 | 0.996 | 0.981 | 1.010 | 1.027 | 1.008 | 1.046 |
| Influenza and pneumonia | 5 | 1.000 | 0.986 | 1.014 | 1.027 | 1.007 | 1.047 |
| Influenza and pneumonia | 6 | 1.022 | 1.009 | 1.035 | 1.049 | 1.028 | 1.070 |
| Chronic respiratory disease | 0 | 0.998 | 0.987 | 1.009 | 0.998 | 0.987 | 1.009 |
| Chronic respiratory disease | 1 | 1.017 | 1.004 | 1.030 | 1.015 | 1.002 | 1.028 |
| Chronic respiratory disease | 2 | 0.995 | 0.983 | 1.008 | 1.010 | 0.996 | 1.025 |
| Chronic respiratory disease | 3 | 1.008 | 0.996 | 1.022 | 1.019 | 1.003 | 1.034 |
| Chronic respiratory disease | 4 | 0.994 | 0.981 | 1.006 | 1.012 | 0.996 | 1.029 |
| Chronic respiratory disease | 5 | 1.001 | 0.988 | 1.014 | 1.013 | 0.996 | 1.030 |
| Chronic respiratory disease | 6 | 1.012 | 1.000 | 1.024 | 1.025 | 1.007 | 1.043 |
| Diseases of esophagus, stomach and duodenum | 0 | 0.987 | 0.960 | 1.014 | 0.987 | 0.960 | 1.014 |
| Diseases of esophagus, stomach and duodenum | 1 | 1.034 | 1.003 | 1.066 | 1.020 | 0.989 | 1.052 |
| Diseases of esophagus, stomach and duodenum | 2 | 0.997 | 0.967 | 1.028 | 1.017 | 0.983 | 1.053 |
| Diseases of esophagus, stomach and duodenum | 3 | 0.990 | 0.959 | 1.022 | 1.007 | 0.971 | 1.045 |
| Diseases of esophagus, stomach and duodenum | 4 | 0.995 | 0.964 | 1.026 | 1.002 | 0.963 | 1.042 |
| Diseases of esophagus, stomach and duodenum | 5 | 1.009 | 0.978 | 1.041 | 1.011 | 0.970 | 1.054 |
| Diseases of esophagus, stomach and duodenum | 6 | 0.996 | 0.968 | 1.024 | 1.007 | 0.965 | 1.050 |
| Diseases of liver | 0 | 1.003 | 0.993 | 1.013 | 1.003 | 0.993 | 1.013 |
| Diseases of liver | 1 | 1.000 | 0.989 | 1.012 | 1.003 | 0.992 | 1.014 |
| Diseases of liver | 2 | 1.006 | 0.995 | 1.017 | 1.009 | 0.997 | 1.022 |
| Diseases of liver | 3 | 0.995 | 0.984 | 1.006 | 1.004 | 0.991 | 1.018 |
| Diseases of liver | 4 | 1.003 | 0.992 | 1.014 | 1.007 | 0.993 | 1.022 |
| Diseases of liver | 5 | 1.008 | 0.997 | 1.019 | 1.015 | 1.000 | 1.031 |
| Diseases of liver | 6 | 1.003 | 0.993 | 1.013 | 1.018 | 1.003 | 1.034 |
| Disorders of gallbladder, biliary tract and pancreas | 0 | 1.012 | 0.987 | 1.038 | 1.012 | 0.987 | 1.038 |
| Disorders of gallbladder, biliary tract and pancreas | 1 | 0.995 | 0.967 | 1.024 | 1.007 | 0.980 | 1.036 |
| Disorders of gallbladder, biliary tract and pancreas | 2 | 1.020 | 0.992 | 1.049 | 1.027 | 0.996 | 1.059 |
| Disorders of gallbladder, biliary tract and pancreas | 3 | 0.992 | 0.964 | 1.021 | 1.019 | 0.986 | 1.053 |
| Disorders of gallbladder, biliary tract and pancreas | 4 | 1.008 | 0.980 | 1.036 | 1.027 | 0.992 | 1.063 |
| Disorders of gallbladder, biliary tract and pancreas | 5 | 0.989 | 0.962 | 1.018 | 1.016 | 0.979 | 1.054 |
| Disorders of gallbladder, biliary tract and pancreas | 6 | 1.000 | 0.975 | 1.025 | 1.016 | 0.979 | 1.054 |
| Renal failure | 0 | 1.001 | 0.983 | 1.018 | 1.001 | 0.983 | 1.018 |
| Renal failure | 1 | 1.005 | 0.986 | 1.025 | 1.006 | 0.986 | 1.025 |
| Renal failure | 2 | 0.987 | 0.968 | 1.006 | 0.992 | 0.971 | 1.013 |
| Renal failure | 3 | 1.014 | 0.994 | 1.034 | 1.006 | 0.983 | 1.029 |
| Renal failure | 4 | 1.008 | 0.988 | 1.028 | 1.013 | 0.989 | 1.039 |
| Renal failure | 5 | 1.009 | 0.990 | 1.029 | 1.023 | 0.997 | 1.050 |
| Renal failure | 6 | 1.012 | 0.994 | 1.030 | 1.035 | 1.008 | 1.062 |
| Suicide | 0 | 1.021 | 0.989 | 1.054 | 1.021 | 0.989 | 1.054 |
| Suicide | 1 | 0.979 | 0.945 | 1.015 | 1.000 | 0.965 | 1.035 |
| Suicide | 2 | 1.018 | 0.982 | 1.055 | 1.018 | 0.979 | 1.058 |
| Suicide | 3 | 0.985 | 0.950 | 1.022 | 1.002 | 0.961 | 1.045 |
| Suicide | 4 | 0.989 | 0.953 | 1.026 | 0.991 | 0.948 | 1.036 |
| Suicide | 5 | 1.000 | 0.965 | 1.037 | 0.991 | 0.946 | 1.039 |
| Suicide | 6 | 1.021 | 0.989 | 1.054 | 1.013 | 0.966 | 1.062 |
